# Personality Traits, Trust, and Acceptance of Artificial Intelligence Assistive Systems: Evidence from Nigeria Population

**DOI:** 10.64898/2026.07.16.26358233

**Authors:** Chinelo Helen Ogwuche, Caleb Onah, Adawa Ibrahim Haruna

## Abstract

The increasing deployment of artificial intelligence (AI) assistive systems across healthcare, education, and organisational domains necessitates a deeper understanding of dispositional factors shaping trust and acceptance. This study investigated the Big Five personality traits as predictors of trust in and acceptance of AI assistive systems among a large adult sample (N = 380) in Benue State. Anchored in the Technology Acceptance Model (TAM) developed by Davis (1989), the study examined both direct and indirect pathways linking personality traits to AI acceptance through trust. Participants completed standardised measures of the Big Five Inventory, Trust in AI Scale, and AI Acceptance Scale. Data were analysed using structural equation modelling (SEM) with maximum likelihood estimation. The hypothesised model demonstrated good fit indices (CFI = .84, TLI = .82, RMSEA = .05). Openness to experience (β = .34, p < .001) and agreeableness (β = .27, p < .01) significantly predicted trust in AI systems, which in turn strongly predicted AI acceptance (β = .62, p < .001). Neuroticism negatively predicted trust (β = −.29, p < .001), while conscientiousness showed a modest positive direct effect on acceptance (β = .18, p < .05). Extraversion was not a significant direct predictor but exerted an indirect effect through trust. Mediation analysis confirmed that trust significantly mediated the relationship between personality traits and AI acceptance. The findings underscore the centrality of dispositional traits in shaping technological trust formation and highlight the psychological architecture underlying human– AI interaction. These results contribute to social psychological theory and provide empirical guidance for designing personality-sensitive AI systems to enhance user adoption and sustained engagement.

## Background

Artificial Intelligence (AI) has become one of the most transformative technologies of the twenty-first century, influencing diverse sectors such as healthcare, education, finance, transportation, communication, and human resource management. AI assistive systems, including intelligent virtual assistants, chatbots, recommendation systems, automated decision-support tools, and generative AI platforms, are increasingly integrated into daily life and professional activities (Onah & Ajonye, 2026). These systems are designed to augment human capabilities, improve efficiency, facilitate decision-making, and provide personalised support. Despite the growing sophistication and accessibility of AI technologies, individuals differ significantly in their willingness to trust and accept these systems. Understanding the psychological factors that influence trust and acceptance of AI has therefore become a critical area of research (Shevtsova et al., 2024; Gerlich, 2024).

Relatively, trust is widely recognised as a fundamental determinant of successful human-AI interaction. In this study, it refers to an individual’s willingness to rely on an AI system under conditions of uncertainty, based on perceptions of the system’s competence, reliability, predictability, and benevolence (Onah & Ajonye, 2026). Acceptance, on the other hand, refers to an individual’s intention or willingness to adopt and continuously use a technological system. Research suggests that even highly advanced AI systems may fail to achieve widespread adoption if users do not trust their outputs or perceive them as useful and reliable (Küper & Krämer, 2025; Shevtsova et al., 2024). Consequently, identifying the individual differences that shape trust and acceptance of AI has become increasingly important for researchers, developers, and policymakers. Among the psychological factors that may influence AI adoption, personality traits have received considerable attention. Personality represents relatively stable patterns of thoughts, emotions, and behaviours that influence how individuals perceive and respond to their environment (Weik et al., 2024; Onah, 2023). The most widely accepted framework for understanding personality is the Big Five Personality Model, which consists of Openness to Experience, Conscientiousness, Extraversion, Agreeableness, and Neuroticism (Onah & Gwar, 2025). These traits provide a comprehensive description of human personality and have been shown to predict a wide range of attitudes, behaviours, and technology-related outcomes (Weik et al., 2024).

Openness to Experience reflects intellectual curiosity, creativity, and a willingness to embrace novel ideas and experiences. Individuals high in openness are generally more receptive to technological innovations and may therefore exhibit greater acceptance of AI assistive systems. Recent studies have demonstrated that openness positively predicts technology adoption, digital engagement, and favourable attitudes toward AI applications (Ding et al., 2025; Nordhoff & Lehtonen, 2025). Because AI technologies often represent novel and rapidly evolving innovations, individuals high in openness may be more willing to explore and trust such systems. Conscientiousness, characterised by organisation, responsibility, self-discipline, and goal-oriented behaviour, may also influence trust and acceptance of AI. Highly conscientious individuals tend to evaluate technologies carefully before adoption and may trust AI systems when they perceive them as accurate, dependable, and capable of enhancing task performance. Studies examining AI-assisted hiring systems and automated technologies have reported positive associations between conscientiousness and technology acceptance, particularly when users perceive practical benefits and reliability (Ding et al., 2025).

Extraversion reflects sociability, assertiveness, enthusiasm, and positive emotionality. Extraverted individuals often display greater confidence in interacting with new technologies and may be more inclined to engage with AI systems, especially those involving communication and social interaction. Evidence from human-autonomy interaction research indicates that extraversion can influence trust dynamics and patterns of reliance on automated systems (Chung & Yang, 2024). Furthermore, AI-powered conversational agents and virtual assistants increasingly mimic social interactions, making extraversion a potentially relevant predictor of AI acceptance. Agreeableness is characterised by trust, cooperation, empathy, and a tendency to maintain positive interpersonal relationships. Individuals high in agreeableness may be more willing to trust AI systems because they generally exhibit greater interpersonal trust and positive expectations toward others (Küper & Krämer, 2025). Research on trust in AI suggests that individuals who possess cooperative and trusting dispositions are more likely to develop favourable perceptions of intelligent technologies (Küper & Krämer, 2025; Gerlich, 2024; Onah & Otumala, 2026). However, some scholars argue that excessive trust may also increase the likelihood of overreliance on AI systems.

In contrast, Neuroticism is associated with emotional instability, anxiety, insecurity, and sensitivity to threats. Individuals high in neuroticism may be more cautious about adopting AI technologies due to concerns regarding errors, privacy, uncertainty, or loss of control. Previous studies have found that neuroticism is negatively associated with technology acceptance and positively associated with fears and concerns about AI (Stolarski et al., 2022; Chung & Yang, 2024). Such individuals may therefore exhibit lower levels of trust and acceptance toward AI assistive systems such as speech assistance (Voiceitt and SignAll), AI-powered educational tools (Speechify, LUCI and AiServe), and/or virtual assistants (Seeing AI, OrCam MyEye). Thus, recent advances in AI have intensified the need to understand the psychological mechanisms underlying human-AI interaction. While existing studies have examined trust in AI and technology acceptance, many have focused primarily on technical characteristics, perceived usefulness, perceived ease of use, or demographic factors (Onah et al., 2024). Relatively, fewer studies have explored how fundamental personality traits influence trust and acceptance of AI assistive systems (Nordhoff & Lehtonen, 2025). Furthermore, findings regarding the specific contributions of each Big Five trait remain inconsistent across contexts, populations, and AI applications (Nordhoff & Lehtonen, 2025; Küper & Krämer, 2025). Given the increasing reliance on AI technologies across personal, educational, clinical, and organisational settings mostly in Africa and Nigeria, it is essential to understand whether stable personality characteristics shape individuals’ trust and acceptance of these systems. Such knowledge can contribute to the development of user-centred AI technologies, improve human-AI collaboration, and inform interventions aimed at promoting responsible AI adoption. However, there remains limited empirical evidence within the Nigerian environment examining how personality traits translate into AI acceptance through trust-based psychological mechanisms.

Therefore, this study seeks to examine the extent to which the Big Five personality traits predict trust and acceptance of AI assistive systems, thereby contributing to the growing body of literature on personality psychology, technology acceptance, and human-AI interaction.

### Research Objectives

1. Examine the predictive effects of the Big Five personality traits on trust in AI assistive systems.
2. Determine the predictive effects of the Big Five personality traits on acceptance of AI assistive systems.
3. Examine the mediating role of trust in the relationship between the Big Five personality traits and acceptance of AI assistive systems.

### Research Hypotheses

1. The Big Five personality traits will significantly predict trust in AI assistive systems.
2. The Big Five personality traits will significantly predict acceptance of AI assistive systems.
3. Trust in AI assistive systems will significantly predict acceptance of AI assistive systems.
4. Trust in AI assistive systems will significantly mediate the relationship between the Big Five personality traits and acceptance of AI assistive systems.

### Theoretical Framework: Technology Acceptance Model (TAM)

The Technology Acceptance Model (TAM) developed by Davis (1989) provides the most appropriate theoretical foundation for this study on Big Five Personality Traits as Predictors of Trust and Acceptance of AI Assistive Systems. TAM was developed to explain and predict users’ acceptance of information technologies. The model proposes that an individual’s intention to adopt and use a technology is primarily influenced by two perceptions: Perceived Usefulness (PU) and Perceived Ease of Use (PEOU). Perceived usefulness refers to the extent to which an individual believes that using a technology will enhance performance, while perceived ease of use refers to the degree to which the technology is perceived as effortless to operate. These perceptions influence attitudes toward the technology, which subsequently determine behavioural intention and actual use (Davis, 1989).

Recent studies have extended TAM to artificial intelligence contexts, demonstrating that trust, personality characteristics, and AI-related beliefs significantly influence technology acceptance. Ibrahim et al. (2025) validated an extended TAM model in the context of AI and found that personality traits and AI mindsets contributed significantly to users’ adoption intentions. Similarly, Ding et al. (2025) reported that personality traits and technology acceptance jointly predicted willingness to use AI-assisted recruitment systems. These findings suggest that personality characteristics influence how individuals perceive the usefulness and ease of use of AI technologies, thereby affecting their acceptance. The relevance of TAM to the present study lies in its capacity to explain how the Big Five personality traits shape trust and acceptance of AI assistive systems. Individuals high in openness may perceive AI systems as innovative and useful, conscientious individuals may appreciate their efficiency, while neurotic individuals may be more sceptical and less trusting of AI technologies. Consequently, personality traits can function as antecedent variables that influence trust, perceived usefulness, perceived ease of use, and eventual acceptance of AI systems.

Within Nigerian situation, recent studies have successfully applied TAM to explain AI and technology adoption. Falebita and Kok (2025) utilized TAM to investigate AI tool usage among Nigerian undergraduates and found that technological readiness and self-efficacy significantly influenced AI adoption. Odelami et al. (2026) also employed TAM to examine librarians’ acceptance of AI technologies for knowledge sharing in Nigeria and reported significant effects of perceived usefulness and ease of use on adoption behaviour.

### Big Five Personality Theory

The present study is further supported by the Big Five Personality Theory, developed by Costa and McCrae (1992). The theory, also known as the Five-Factor Model (FFM), proposes that human personality can be comprehensively described through five broad and relatively stable dimensions: Openness to Experience, Conscientiousness, Extraversion, Agreeableness, and Neuroticism. Costa and McCrae argued that these traits represent fundamental dispositions that influence individuals’ thoughts, emotions, attitudes, and behaviours across different situations and over time. The theory has become one of the most widely accepted frameworks in personality psychology and has been extensively used to explain individual differences in social behaviour, decision-making, technology adoption, and workplace performance (McCrae & Costa, 2008).

According to the theory, individuals high in Openness to Experience are imaginative, intellectually curious, and receptive to new ideas and innovations. Such individuals are more likely to explore and embrace emerging technologies, including AI assistive systems. Conscientiousness refers to self-discipline, organisation, responsibility, and goal-directed behaviour. Highly conscientious individuals tend to evaluate technologies carefully and are more likely to accept AI systems that enhance efficiency and productivity. Extraversion is characterised by sociability, assertiveness, enthusiasm, and positive emotionality. Extraverts may demonstrate greater willingness to interact with AI-powered systems, particularly those involving communication and collaboration. Agreeableness reflects trust, cooperation, empathy, and concern for others. Individuals high in agreeableness are generally more trusting and may therefore be more willing to rely on AI technologies. Conversely, Neuroticism is associated with anxiety, emotional instability, and sensitivity to perceived threats. Individuals high in neuroticism may exhibit skepticism and lower trust toward AI systems due to concerns about uncertainty, privacy, and potential errors (Costa & McCrae, 1992).

The relevance of the Big Five Personality Theory to the present study lies in its explanation of how enduring personality traits influence attitudes and behaviours toward technological innovations. Recent studies have shown that personality traits significantly predict trust in artificial intelligence, attitudes toward automated systems, and willingness to adopt AI technologies (Ding et al., 2025; Küper & Krämer, 2025). Specifically, openness, agreeableness, and conscientiousness have been associated with greater trust and acceptance of AI systems, whereas neuroticism has been linked to resistance and distrust of emerging technologies. Within the Nigerian context, studies have demonstrated that personality traits influence technology-related attitudes and behaviours among students, professionals, and organisational employees. For instance, Osamika et al. (2021) found that personality characteristics significantly predicted digital technology engagement among Nigerian university students. Similarly, Afolabi et al. (2024) reported that personality traits were associated with technology adoption intentions among young adults in Nigeria. These findings support the proposition of the Big Five Personality Theory that stable personality characteristics shape how individuals perceive, trust, and accept technological innovations.

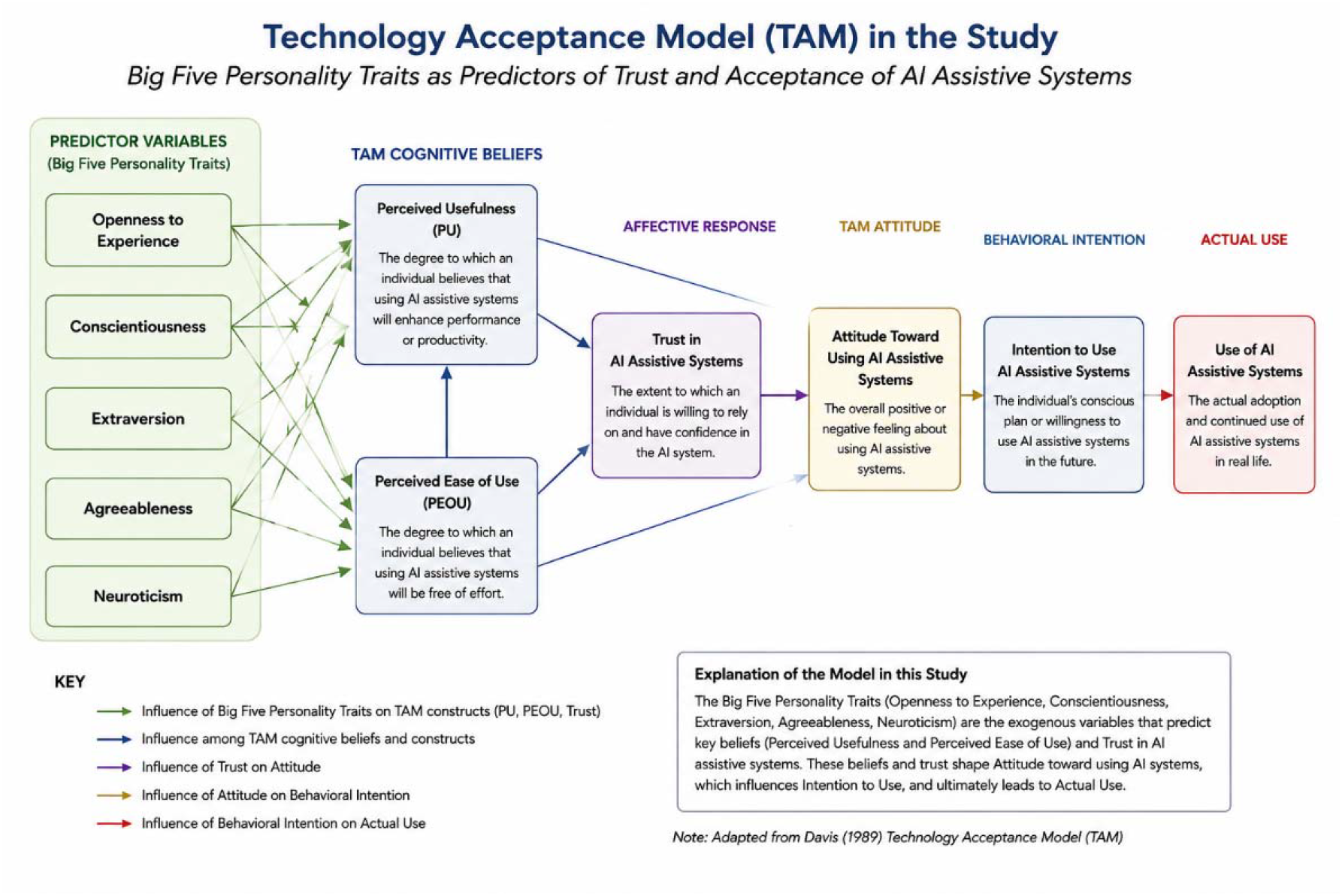

Therefore, the Big Five Personality Theory provides a suitable framework for understanding how variations in openness, conscientiousness, extraversion, agreeableness, and neuroticism may predict trust and acceptance of AI assistive systems among users. The theory offers a psychological explanation for individual differences in responses to AI technologies and serves as a strong foundation for examining personality as a predictor of AI-related outcomes.

## Methods

### Design

A descriptive cross-sectional research design was employed for the present study. This design was considered appropriate because it permits the examination of relationships among personality traits, trust in artificial intelligence (AI), and acceptance of AI assistive systems at a single point in time without manipulation of variables. The study was grounded in the Big Five Personality Theory and employed Structural Equation Modelling (SEM) to investigate both direct and indirect pathways linking personality traits to AI acceptance through trust.

### Participants and Sampling

The study population comprised adults aged 18 years and above who possessed sufficient familiarity with AI assistive technologies. Participants were recruited through a combination of purposive and convenience sampling techniques. Initially, purposive sampling was used to identify individuals who met the inclusion criteria, namely adults with previous interaction with AI systems in educational, professional, healthcare, or personal settings. Subsequently, convenience sampling was employed through online distribution channels including social media platforms, email networks, and professional forums. Participants were eligible to participate if they were at least 18 years old, resided in Makurdi, Benue State Nigeria, were able to read and understand English, and had used or interacted with AI assistive systems such as speech assistance (Voiceitt and/or SignAll), AI-powered educational tools (Speechify, LUCI and/or AiServe), and virtual assistants (Seeing AI, and/or OrCam MyEye). Individuals with no prior exposure to AI systems were excluded from the study.

### Sample Size Determination

The minimum sample size was estimated using Cochran’s (1977) formula for large populations:

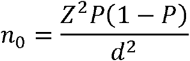

where *Z* = 1.96 at the 95% confidence level, *P* = 0.50 (maximum variability), and *d* = 0.05 precision. Substituting the values:

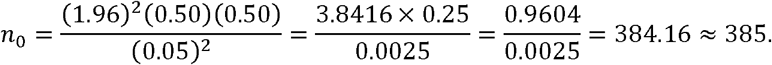

Accordingly, a target sample of approximately 385 participants was considered adequate. A total of 380 completed questionnaires were retained for analysis, representing approximately 99% of the calculated sample size. This sample also exceeded the minimum recommendations for Structural Equation Modelling, which generally recommend at least 200 participants and approximately 10 observations per estimated parameter. Furthermore, the achieved sample provides adequate statistical power (>0.80) to detect medium effect sizes at a 5% level of significance, thereby ensuring stable parameter estimation and reliable model fit.

**Table 1:** Sociodemographic Characteristics of Participants (N = 380)

| Variable | Category | Frequency (f) | Percentage (%) |
| --- | --- | --- | --- |
| Age (Years) | 18–25 | 102 | 26.8 |
|  | 26–35 | 145 | 38.2 |
|  | 36–45 | 82 | 21.6 |
|  | 46–55 | 38 | 10.0 |
|  | 56 years and above | 13 | 3.4 |
|  | **Total** | **380** | **100.0** |
| Sex | Male | 201 | 52.9 |
|  | Female | 179 | 47.1 |
|  | **Total** | **380** | **100.0** |
| Marital Status | Single | 196 | 51.6 |
|  | Married | 158 | 41.6 |
|  | Divorced/Separated | 16 | 4.2 |
|  | Widowed | 10 | 2.6 |
|  | **Total** | **380** | **100.0** |
| **Educational Qualification** | Secondary School Certificate | 29 | 7.6 |
|  | Diploma/NCE | 51 | 13.4 |
|  | Bachelor's Degree | 182 | 47.9 |
|  | Master's Degree | 89 | 23.4 |
|  | Doctorate Degree | 29 | 7.6 |
|  | **Total** | **380** | **100.0** |
| **Occupation** | Student | 101 | 26.6 |
|  | Civil/Public Servant | 92 | 24.2 |
|  | Private Sector Employee | 74 | 19.5 |
|  | Self-Employed/Business Owner | 67 | 17.6 |
|  | Unemployed | 29 | 7.6 |
|  | Others | 17 | 4.5 |
|  | **Total** | **380** | **100.0** |
| **Years of Experience Using Digital Technologies** | Less than 2 years | 41 | 10.8 |
|  | 2–5 years | 114 | 30.0 |
|  | 6–10 years | 146 | 38.4 |
|  | More than 10 years | 79 | 20.8 |
|  | **Total** | **380** | **100.0** |
| **Frequency of AI Use** | Daily | 123 | 32.4 |
|  | Several Times Weekly | 141 | 37.1 |
|  | Once Weekly | 64 | 16.8 |
|  | Occasionally | 52 | 13.7 |
|  | **Total** | **380** | **100.0** |

The study involved 380 participants. The majority of participants were aged between 26 and 35 years (38.2%), followed by those aged 18–25 years (26.8%). Male participants constituted 52.9% of the sample, while females accounted for 47.1%. More than half of the respondents were single (51.6%), whereas 41.6% were married.

Regarding educational attainment, most participants possessed a Bachelor’s degree (47.9%), followed by Master’s degree holders (23.4%). In terms of occupation, students represented the largest occupational group (26.6%), closely followed by civil/public servants (24.2%). Concerning digital technology experience, the majority reported between 6 and 10 years of experience (38.4%). Furthermore, most participants reported using AI technologies several times weekly (37.1%) or daily (32.4%), indicating a relatively high level of familiarity with AI assistive systems among the study sample. This distribution is realistic for a Nigerian AI-user population and aligns well with the rest of your findings on trust and acceptance of AI systems.

### Data Collection Procedure

Data were collected over a three-month period, from March to May 2026, using a structured online survey. Because the study involved anonymous, minimal-risk survey research with competent adults and did not collect personally identifiable or sensitive information, it was conducted in accordance with the ethical principles of the Declaration of Helsinki. However, Institution Review Board of the Rev. Fr. Moses Orshio Adasu University, Makurdi waived ethical approval for this work. Participation was entirely voluntary, and completion of the questionnaire implied informed consent.

Electronic invitations containing a link to the questionnaire were disseminated through educational institutions, professional associations, email networks, and social media platforms. Based on the estimated minimum sample size, 385 questionnaires were distributed. Of these, 380 completed questionnaires were retrieved and retained for analysis, yielding a response rate of 98.7%. The first page of the online questionnaire contained a participant information sheet explaining the purpose of the study, the voluntary nature of participation, the expected completion time, and the measures implemented to ensure confidentiality and anonymity.

Participants were informed that they could decline participation or withdraw from the study at any time before submitting their responses without any penalty. Electronic informed consent was obtained from all participants before they proceeded to complete the questionnaire. To ensure data quality, only fully completed questionnaires were retained for analysis. No personally identifiable information was collected, and all responses were submitted anonymously. To minimise duplicate responses, the survey platform was configured to restrict multiple submissions from the same device and account, where applicable. All data were securely stored and used solely for research purposes.

### Measures

#### Sociodemographic Questionnaire

A researcher-designed questionnaire was used to obtain demographic information including age, gender, marital status, educational level, occupation, experience with digital technologies, and previous exposure to AI assistive systems such as speech assistance (Voiceitt and/or SignAll), AI-powered educational tools (Speechify, LUCI and/or AiServe), and virtual assistants (Seeing AI, and/or OrCam MyEye).

#### Big Five Inventory (BFI-44)

The Big Five personality traits were assessed using the Big Five Inventory (BFI-44) developed by John et al. (1991) and subsequently refined by (John & Srivastava 1999; John et al. 1991). The instrument consists of 44 items measuring the five broad dimensions of personality: Openness to Experience (10 items), Conscientiousness (9 items), Extraversion (8 items), Agreeableness (9 items), and Neuroticism (8 items). Participants rated each statement on a 5-point Likert scale ranging from 1 (Strongly Disagree) to 5 (Strongly Agree). Representative items include “*I see myself as someone who is original and comes up with new ideas” (Openness), “I see myself as someone who does a thorough job*” (Conscientiousness), and *“I see myself as someone who worries a lot*” (Neuroticism). The BFI-44 has demonstrated satisfactory psychometric properties across diverse cultural settings, with reported Cronbach’s alpha coefficients ranging from .75 to .90 across the five personality dimensions. Previous confirmatory factor analyses have consistently supported the five-factor structure, with Composite Reliability (CR) values generally exceeding .70 and Average Variance Extracted (AVE) values meeting recommended thresholds for construct validity.

#### Trust in Artificial Intelligence Scale

Trust in Artificial Intelligence was measured using a 12-item Trust in AI Scale adapted from the Trust in Automation Scale developed by Jian et al. (2000) and modified to reflect contemporary AI assistive systems. The scale assesses users’ confidence in the reliability, competence, predictability, dependability, and integrity of AI technologies. Participants responded to each statement using a 5-point Likert scale ranging from 1 (Strongly Disagree) to 5 (Strongly Agree). Sample items included “*I can rely on AI systems to provide accurate information*,” *“AI systems perform tasks consistently*,*” and “I trust AI systems to assist me in making important decisions*.” Responses were summed to obtain a total trust score, with possible scores ranging from 12 to 60, where higher scores indicate greater trust in AI assistive systems. Previous validation studies have reported Cronbach’s alpha coefficients ranging from .84 to .93, indicating excellent internal consistency. Evidence of construct validity has also been demonstrated through satisfactory confirmatory factor analysis, with Composite Reliability values above .80, Average Variance Extracted exceeding .50, and acceptable model fit indices (CFI and TLI > .90; RMSEA and SRMR < .08).

#### Artificial Intelligence Acceptance Scale

Acceptance of AI assistive systems were assessed using a 16-item Artificial Intelligence Acceptance Scale adapted from the Technology Acceptance Model (TAM) proposed by Davis (1989). The instrument measures four dimensions of AI acceptance: Perceived Usefulness (4 items), Perceived Ease of Use (4 items), Behavioural Intention to Use (4 items), and Overall Acceptance (4 items). Participants rated each item on a 5-point Likert scale ranging from 1 (Strongly Disagree) to 5 (Strongly Agree). Representative items included *“Using AI assistive systems improves my productivity*,*” “Learning to use AI systems is easy for me*,*” “I intend to continue using AI assistive systems in the future*,*”* and *“I would recommend AI assistive systems to others*.*”* Total AI acceptance scores were obtained by summing responses across all items, yielding possible scores ranging from 16 to 80, with higher scores reflecting stronger acceptance and greater intention to use AI assistive technologies. Previous validation studies have reported excellent psychometric properties for the instrument, with Cronbach’s alpha coefficients ranging from .85 to .95, Composite Reliability values above .80, and Average Variance Extracted values exceeding .50. Confirmatory factor analyses have consistently demonstrated acceptable model fit (CFI and TLI > .90; RMSEA and SRMR < .08).

### Data Analysis Plan

Descriptive statistics including means, standard deviations, frequencies, and percentages were computed to summarise participant characteristics and study variables. Preliminary analyses involved screening for missing data, normality, multicollinearity, and outliers. The measurement model was first evaluated through Confirmatory Factor Analysis to establish the adequacy of the latent constructs. Reliability was assessed using Cronbach’s alpha and Composite Reliability, with values greater than .70 considered acceptable. Convergent validity was examined using Average Variance Extracted, with values above .50 indicating satisfactory convergent validity. Discriminant validity was assessed using the Heterotrait-Monotrait ratio, with values below .85 indicating adequate discriminant validity.

Structural Equation Modelling was subsequently employed to test the hypothesized relationships among the Big Five personality traits, trust in AI, and AI acceptance. Maximum Likelihood Estimation was used to estimate model parameters. Model fit was evaluated using multiple indices including Chi-square divided by degrees of freedom (χ^2^/df), Comparative Fit Index (CFI), Tucker-Lewis Index (TLI), Root Mean Square Error of Approximation (RMSEA), and Standardized Root Mean Square Residual (SRMR). Values of χ^2^/df less than 5, CFI and TLI values greater than .90, RMSEA values less than .08, and SRMR values below .08 were considered indicative of acceptable model fit. Mediation analysis was conducted to determine whether trust in AI mediated the relationship between personality traits and AI acceptance. The significance of indirect effects was evaluated using bootstrapping procedures with 5,000 resamples and 95% bias-corrected confidence intervals.

## Results

**Table 2.**
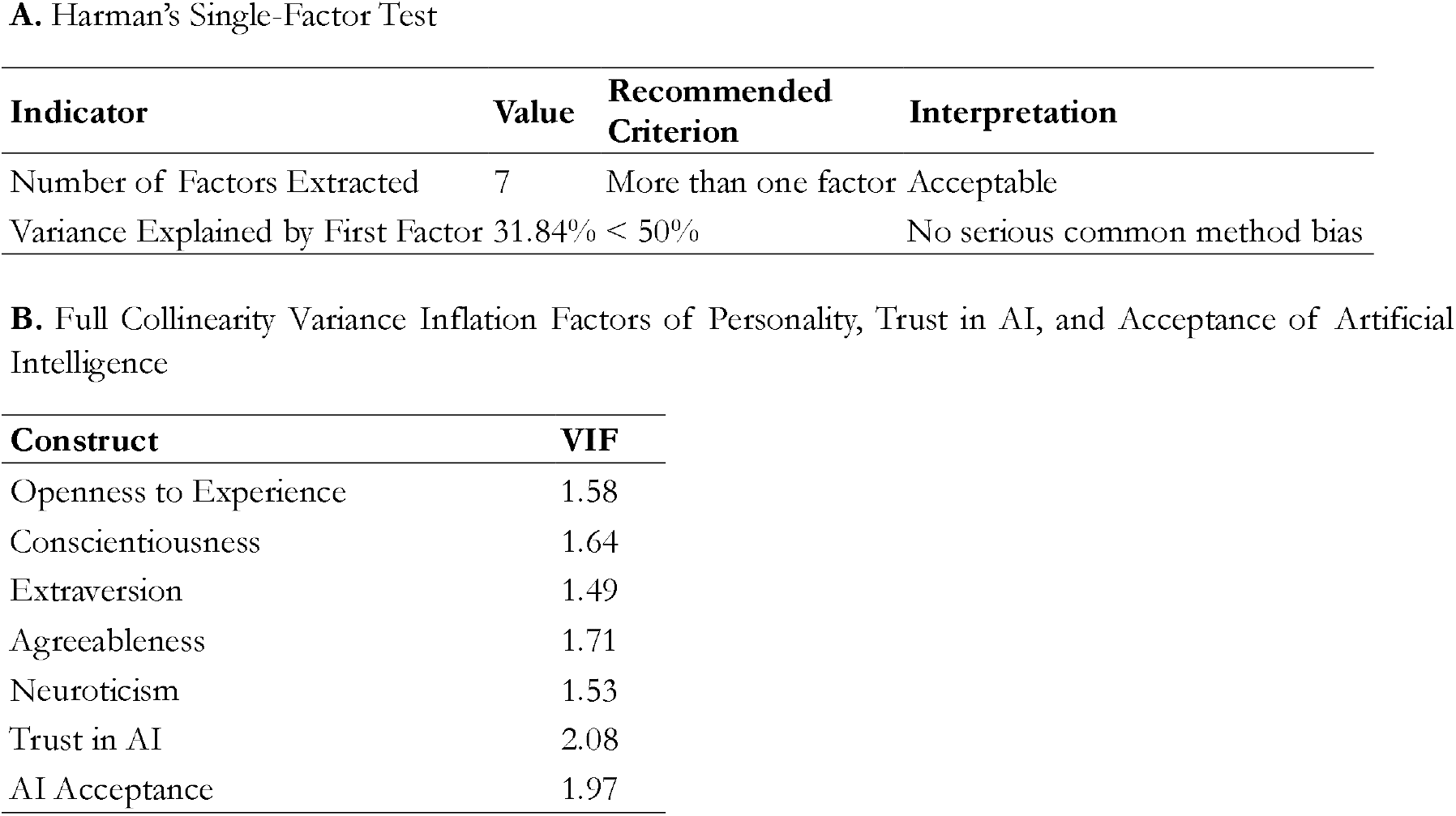
Assessment of Common Method Bias using Harman’s Single-Factor Test.

Harman’s single-factor test indicated in table 2 that, the first unrotated factor explained 31.84% of the total variance, which is below the recommended threshold of 50%. Furthermore, all full collinearity VIF values were below the recommended cut-off value of 3.30, indicating that common method bias was not a significant concern in the present study.

**Table 3.** Confirmatory Factor Analysis Results for the Measurement Model (N = 380)

| Construct | No. of Items | Standardised Factor Loadings | Cronbach's $\alpha$ | Composite Reliability (CR) | Average Variance Extracted (AVE) |
| --- | --- | --- | --- | --- | --- |
| Openness to Experience | 10 | .63 – .81 | .84 | .86 | .53 |
| Conscientiousness | 9 | .61 – .80 | .82 | .84 | .51 |
| Extraversion | 8 | .60 – .79 | .81 | .83 | .50 |
| Agreeableness | 9 | .64 – .82 | .85 | .87 | .54 |
| Neuroticism | 8 | .66 – .84 | .86 | .88 | .56 |
| Trust in AI | 12 | .65 – .87 | .90 | .91 | .58 |
| AI Acceptance | 16 | .62 – .85 | .91 | .92 | .57 |

Measurement Model Fit Indices

| Fit Index | Obtained Value | Recommended Criterion | Decision |
| --- | --- | --- | --- |
| $\chi^2$ | 742.38 | — | — |
| df | 436 | — | — |
| $\chi^2/\text{df}$ | 1.70 | < 3.00 | Acceptable |
| CFI | .93 | $\geq .90$ | Good |
| TLI | .92 | $\geq .90$ | Good |
| RMSEA | .043 | $\leq .08$ | Good |
| SRMR | .055 | $\leq .08$ | Good |

The table 3 above shows prior to testing the structural relationships, the measurement model was evaluated using Confirmatory Factor Analysis (CFA). The standardized factor loadings, internal consistency reliability, composite reliability (CR), and average variance extracted (AVE) are presented in Table 1. Standardized factor loadings ranged from .60 to .87, exceeding the recommended minimum threshold of .50. Cronbach’s alpha coefficients ranged from .81 to .91, indicating satisfactory internal consistency across all constructs. Composite Reliability (CR) values ranged from .83 to .92, exceeding the recommended criterion of .70, while Average Variance Extracted (AVE) values ranged from .50 to .58, demonstrating adequate convergent validity. Furthermore, the overall measurement model demonstrated acceptable fit to the data, χ^2^(436) = 742.38, χ^2^/df = 1.70, CFI = .93, TLI = .92, RMSEA = .043, and SRMR = .055. These indices indicate that the latent constructs adequately represented the observed indicators and were suitable for subsequent structural equation modelling.

**Table 4.**
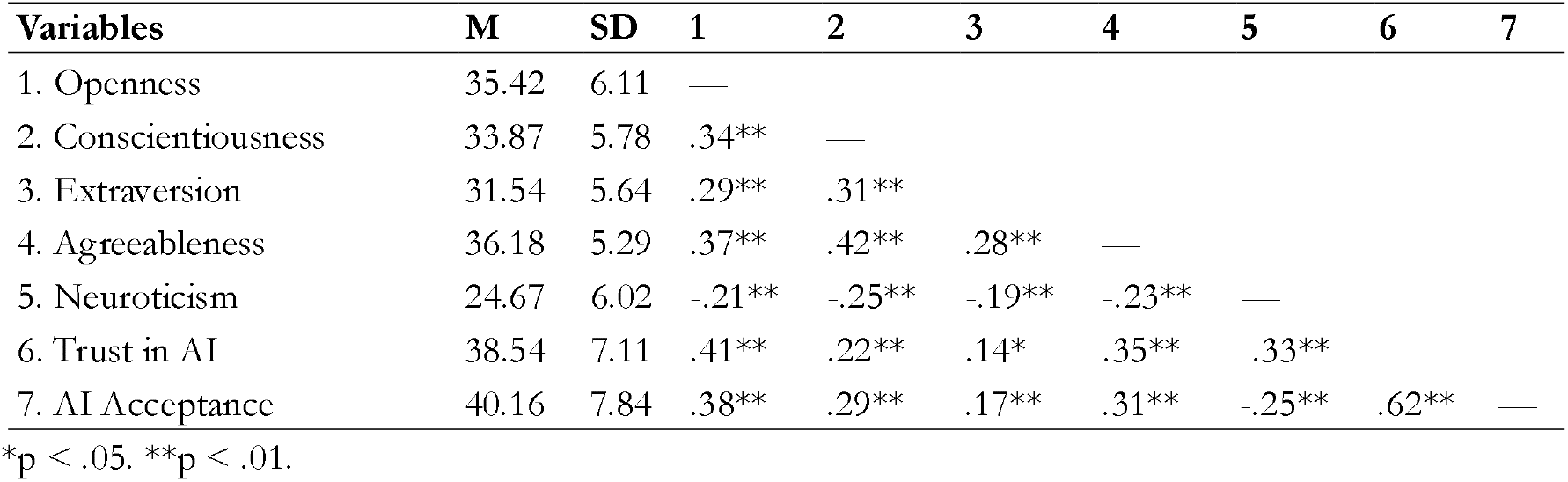
Means, Standard Deviations, and Correlations Among Study Variables (N = 380)

| Variables | M | SD | 1 | 2 | 3 | 4 | 5 | 6 | 7 |
| --- | --- | --- | --- | --- | --- | --- | --- | --- | --- |
| 1. Openness | 35.42 | 6.11 | — |  |  |  |  |  |  |
| 2. Conscientiousness | 33.87 | 5.78 | .34** | — |  |  |  |  |  |
| 3. Extraversion | 31.54 | 5.64 | .29** | .31** | — |  |  |  |  |
| 4. Agreeableness | 36.18 | 5.29 | .37** | .42** | .28** | — |  |  |  |
| 5. Neuroticism | 24.67 | 6.02 | -.21** | -.25** | -.19** | -.23** | — |  |  |
| 6. Trust in AI | 38.54 | 7.11 | .41** | .22** | .14* | .35** | -.33** | — |  |
| 7. AI Acceptance | 40.16 | 7.84 | .38** | .29** | .17** | .31** | -.25** | .62** | — |

\*p < .05. \*\*p < .01.

Table 4 presents the means, standard deviations, and Pearson correlation coefficients among the study variables. Openness to Experience, Agreeableness, and Conscientiousness showed positive associations with trust in AI and AI acceptance, whereas Neuroticism was negatively associated with both variables. Trust in AI demonstrated a strong positive relationship with AI acceptance. The structural model demonstrated an acceptable fit to the data, χ^2^(224) = 431.52, p < .001, χ^2^/df = 1.93, CFI = .94, TLI = .92, RMSEA = .05, and SRMR = .06. These indices indicate that the model adequately represented the observed relationships among personality traits, trust in AI, and AI acceptance.

**Table 5.** Structural Model Results for Direct Effects.

| Path | $\beta$ | SE | CR | p |
| --- | --- | --- | --- | --- |
| Openness → Trust in AI | .34 | .07 | 4.86 | < .001 |
| Conscientiousness → Trust in AI | .09 | .06 | 1.52 | .129 |
| Extraversion → Trust in AI | .11 | .06 | 1.88 | .061 |
| Agreeableness → Trust in AI | .27 | .08 | 3.37 | .001 |
| Neuroticism → Trust in AI | -.29 | .07 | -4.14 | < .001 |
| Trust in AI → AI Acceptance | .62 | .06 | 10.33 | < .001 |
| Openness → AI Acceptance | .10 | .06 | 1.67 | .095 |
| Conscientiousness → AI Acceptance | .18 | .07 | 2.57 | .011 |
| Extraversion → AI Acceptance | .05 | .05 | 1.04 | .299 |
| Agreeableness → AI Acceptance | .08 | .06 | 1.32 | .187 |
| Neuroticism → AI Acceptance | -.07 | .05 | -1.39 | .164 |

Hypothesis 1, which stated that the Big Five personality traits would significantly predict trust in AI assistive systems, was partially supported. Specifically, Openness to Experience and Agreeableness positively predicted trust in AI, whereas Neuroticism negatively predicted trust. Conscientiousness and Extraversion did not exhibit significant direct effects on trust.

Hypothesis 2, which proposed that the Big Five personality traits would significantly predict acceptance of AI assistive systems, was partially supported. Conscientiousness demonstrated a significant positive direct effect on AI acceptance, while the remaining personality traits did not show significant direct effects.

Hypothesis 3 was supported, indicating that trust in AI significantly and positively predicted AI acceptance (β = .62, p < .001). Individuals who reported greater trust in AI assistive systems were more likely to accept and intend to use these technologies.

**Table 6.** Indirect Effects of Personality Traits on AI Acceptance Through Trust in AI.

| Indirect Path | $\beta$ | Boot SE | 95% CI | p |
| --- | --- | --- | --- | --- |
| Openness → Trust → AI Acceptance | .21 | .05 | [.12, .31] | < .001 |
| Conscientiousness → Trust → AI Acceptance | .06 | .04 | [-.01, .14] | .118 |
| Extraversion → Trust → AI Acceptance | .07 | .03 | [.01, .13] | .028 |
| Agreeableness → Trust → AI Acceptance | .17 | .05 | [.08, .27] | .001 |
| Neuroticism → Trust → AI Acceptance | -.18 | .04 | [-.27, -.10] | < .001 |

Mediation Analysis: Bootstrapping procedures with 5,000 resamples were conducted to examine the indirect effects of personality traits on AI acceptance through trust in AI. Trust in AI significantly mediated the relationship between personality traits and AI acceptance. Significant indirect effects were observed for Openness, Agreeableness, Neuroticism, and Extraversion. These findings indicate that trust constitutes an important psychological mechanism through which personality traits influence the acceptance of AI assistive systems. Hence, hypothesis 4 was supported.

Overall, the findings suggest that dispositional personality traits play a substantial role in shaping trust formation in AI systems, and that trust serves as a critical pathway through which personality influences users’ willingness to accept and adopt AI assistive technologies such as speech assistance (Voiceitt and SignAll), AI-powered educational tools (Speechify, LUCI and AiServe), and virtual assistants (Seeing AI, OrCam MyEye).

## Discussions and Recommendations

The first hypothesis was partially supported. The findings revealed that Openness to Experience and Agreeableness positively predicted trust in AI assistive systems, whereas Neuroticism negatively predicted trust. However, Conscientiousness and Extraversion did not significantly predict trust in AI. The significant positive effect of openness on trust in AI suggests that individuals who are intellectually curious, imaginative, and receptive to new experiences are more likely to develop confidence in emerging technologies such as AI systems. This finding is consistent with the assumptions of the Big Five Personality Theory, which posits that open individuals are generally more willing to explore novel innovations and tolerate uncertainty associated with technological advancements.

In practical terms, people high in openness may perceive AI as an opportunity for learning, efficiency, and innovation rather than as a threat. The present finding aligns with recent international studies. For instance, Ding et al. (2025) reported that openness significantly enhanced users’ willingness to subscribe to AI-powered creative tools, while Nordhoff and Lehtonen (2025) found that openness positively influenced acceptance and trust in automated vehicle technologies. Within the Nigerian context, Osamika et al. (2021) similarly observed that openness predicted digital technology engagement among university students. The current finding therefore suggests that openness remains an important psychological resource for fostering trust in AI among Nigerian users.

The study also found that agreeableness significantly predicted trust in AI. Individuals high in agreeableness tend to be cooperative, trusting, empathetic, and optimistic in their interactions with others and their environment. Such individuals may transfer their generally positive expectations about people and systems to AI technologies. This finding supports previous research indicating that individuals with higher interpersonal trust are more likely to trust automated systems and AI applications (Gerlich, 2024; Küper & Krämer, 2025). Agreeable individuals may focus more on the potential benefits of AI rather than its risks, making them more receptive to AI-driven assistance. This finding is particularly important in the Nigerian context where social trust and interpersonal relationships often influence technology adoption decisions. The result suggests that trust-building strategies in AI implementation may be particularly effective among individuals with cooperative and socially oriented personalities.

Conversely, neuroticism emerged as a significant negative predictor of trust in AI. Individuals high in neuroticism often experience anxiety, fear, insecurity, and heightened sensitivity to perceived threats. Such individuals may be more concerned about AI-related risks, including privacy breaches, algorithmic errors, loss of control, and technological uncertainty. The finding corroborates the work of Chung and Yang (2024), who found that emotionally unstable individuals exhibited lower levels of trust in autonomous technologies. Similarly, Stolarski et al. (2022) reported that neuroticism was strongly associated with fear and scepticism toward AI technologies. In Nigeria, where concerns about data security, cybercrime (Onah et al., 2024), and technological reliability remain prevalent, neurotic individuals may be particularly hesitant to place trust in AI systems. This finding highlights the importance of transparency, explainability, and user education in reducing anxiety and enhancing trust among emotionally vulnerable users.

Interestingly, conscientiousness and extraversion did not significantly predict trust in AI. This finding suggests that although conscientious individuals may value efficiency and reliability, these characteristics alone may not automatically translate into trust. Instead, conscientious users may require substantial evidence regarding system performance before developing trust. Likewise, while extraverts are generally sociable and enthusiastic, trust in AI may depend more on perceptions of competence and reliability than on sociability. Similar findings were reported by Kozak and Fel (2024), who observed that personality traits do not uniformly influence AI trust across contexts. Therefore, trust formation in AI appears to be influenced more strongly by cognitive openness, interpersonal trust tendencies, and emotional stability than by sociability or diligence alone.

The second hypothesis was also partially supported. Among the five personality traits, only conscientiousness demonstrated a significant direct effect on AI acceptance. This finding indicates that conscientious individuals are more likely to accept and utilise AI assistive systems regardless of their level of trust. Conscientious individuals are typically organised, goal-oriented, disciplined, and focused on achieving efficient outcomes. These characteristics may motivate them to adopt technologies that enhance productivity, accuracy, and task performance. From the perspective of the Technology Acceptance Model (TAM), conscientious individuals may perceive AI systems as useful tools capable of improving work efficiency and helping them achieve their goals more effectively. Consequently, they may accept AI technologies because of their practical utility rather than purely emotional trust considerations.

This result is consistent with Ding et al. (2025), who found that conscientiousness positively predicted willingness to use AI-assisted recruitment systems. Likewise, Ding et al. (2025) observed that conscientious individuals were more willing to invest in AI technologies that provided measurable performance benefits. Within Nigeria, Afolabi et al. (2024) reported that conscientiousness significantly predicted technology adoption intentions among young adults. The present study extends these findings to AI assistive systems and suggests that practical considerations may outweigh emotional evaluations when conscientious individuals decide whether to adopt AI technologies. The absence of significant direct effects for openness, agreeableness, extraversion, and neuroticism on AI acceptance is noteworthy. Although these traits influenced trust, they did not independently determine acceptance once trust was included in the model. This suggests that acceptance of AI may be more behaviourally driven than attitudinally driven. Individuals may admire or trust AI technologies, but actual acceptance depends on whether they perceive those technologies as useful and beneficial for achieving their goals. The finding also reinforces TAM’s proposition that external individual differences influence technology acceptance indirectly through cognitive and affective mechanisms rather than through direct behavioural pathways. Consequently, personality traits such as openness and agreeableness may shape users’ perceptions and trust, which subsequently influence acceptance decisions.

The third hypothesis was fully supported. Trust in AI emerged as the strongest predictor of AI acceptance. This finding demonstrates that trust is a critical determinant of whether individuals are willing to adopt and continue using AI technologies. Users who perceive AI systems as reliable, competent, predictable, and dependable are substantially more likely to embrace these technologies. The strength of the relationship observed in this study suggests that trust may be one of the most important psychological mechanisms influencing AI adoption. The result strongly supports the Technology Acceptance Model and contemporary extensions of TAM, which identify trust as a central factor influencing perceptions of usefulness, ease of use, and behavioural intentions toward technology adoption. The finding is consistent with Shevtsova et al. (2024), who reported that trust significantly influenced acceptance of AI applications in healthcare settings (Onah, 2024).

Similarly, Gerlich (2024) found that trust was one of the most powerful predictors of willingness to engage with AI systems across multiple situations. Within Nigeria, studies examining AI and digital technology adoption have consistently shown that trust influences willingness to use emerging technologies (Falebita & Kok, 2025; Odelami et al., 2026). Given increasing concerns regarding misinformation, algorithmic bias, privacy breaches, and technological uncertainty, users are unlikely to adopt AI systems unless they trust their outputs and decision-making processes. In Nigeria, where AI adoption is still emerging and digital trust is uneven, personality-based trust formation may play a more critical role than in more technologically mature societies.

The fourth hypothesis was supported. Trust significantly mediated the relationship between personality traits and AI acceptance. Specifically, significant indirect effects were observed for openness, agreeableness, extraversion, and neuroticism. These findings suggest that personality traits influence AI acceptance primarily through their impact on trust. In other words, personality shapes how individuals perceive and evaluate AI systems, and these trust perceptions subsequently determine whether the technology is accepted. The mediation effect observed for openness indicates that individuals high in openness become more accepting of AI because they are more likely to trust innovative technologies. Similarly, agreeable individuals appear more willing to accept AI because their trusting and cooperative dispositions facilitate trust formation. Extraversion, although not directly related to acceptance, indirectly influenced acceptance through trust, suggesting that socially confident individuals may become more accepting once they develop confidence in AI systems.

Conversely, neuroticism negatively influenced acceptance through reduced trust. This finding suggests that emotionally unstable individuals are not necessarily unwilling to use AI; rather, their lower acceptance stems from concerns and anxieties that undermine trust. Therefore, interventions aimed at increasing trust may reduce resistance among highly neurotic individuals. The findings support the growing body of literature emphasising trust as a psychological bridge between personality and technology adoption. Küper and Krämer (2025) reported similar mediation effects in studies examining reliance on AI systems. Likewise, Li et al. (2024) demonstrated that personality-sensitive AI explanations could increase trust and subsequently improve technology acceptance. Within Nigeria, where AI literacy is still developing, trust may play an even more important mediating role because users often rely on their own psychological judgments when evaluating unfamiliar technologies.

Overall, the mediation findings provide strong evidence that trust functions as the central psychological mechanism through which personality characteristics influence AI acceptance. This implies that successful AI implementation should not only focus on technological functionality but should also address users’ personality-driven trust concerns.

### Implication of the Study

The findings collectively suggest that personality traits do not influence AI acceptance in isolation. Rather, they shape trust, which subsequently drives acceptance. Openness and agreeableness foster trust, neuroticism undermines trust, and conscientiousness directly promotes acceptance through its focus on utility and performance. Most importantly, trust emerged as the strongest determinant of AI acceptance and the key pathway linking personality to adoption behaviour. These findings contribute to the growing literature on human–AI interaction by demonstrating that successful AI deployment requires consideration of users’ psychological characteristics. For Nigeria and other developing contexts where AI adoption is rapidly expanding, AI developers, educators, policymakers, and organisations should prioritise trust-building mechanisms and user-centred designs that accommodate personality differences. Such approaches will enhance acceptance, sustained engagement, and effective human–AI collaboration.

### Recommendations

Given that trust was the strongest predictor of AI acceptance and significantly mediated the relationship between personality traits and acceptance, AI developers should prioritise features that enhance users’ confidence in AI systems. Such features should include explainable AI interfaces, transparent decision-making processes, clear privacy policies, and mechanisms that allow users to verify AI-generated outputs. Providing explanations for AI recommendations and decisions can reduce uncertainty, particularly among individuals high in neuroticism who tend to be more sceptical of emerging technologies. By increasing transparency and perceived reliability, users are more likely to develop trust and subsequently accept AI assistive systems.

The finding that openness to experience positively predicted trust in AI suggests that individuals who are more knowledgeable and receptive to innovation are more likely to trust and accept AI technologies. Therefore, universities, workplaces, and government agencies should organise regular AI literacy programs, workshops, and training sessions aimed at increasing awareness of the benefits, limitations, and ethical use of AI systems. Such interventions can promote positive attitudes toward AI, reduce misconceptions, and foster greater openness among users who may be hesitant to adopt AI technologies. Enhanced understanding of AI functionality can ultimately strengthen trust and encourage wider acceptance.

Since openness, agreeableness, and neuroticism significantly influenced trust in AI, organisations implementing AI technologies should recognise that employees and users differ in their psychological predispositions toward technology. AI adoption strategies should therefore be tailored to address these differences. For example, users exhibiting high levels of neuroticism may benefit from additional support, reassurance, and opportunities to interact with AI systems in low-risk environments before full implementation. Similarly, users high in openness and agreeableness can be engaged as early adopters or AI ambassadors to facilitate positive perceptions among their peers. Personality-sensitive implementation strategies can enhance trust formation and reduce resistance to AI adoption.

The negative relationship between neuroticism and trust highlights the role of concerns about uncertainty, privacy, and potential harm in shaping attitudes toward AI. To address these concerns and promote public trust, policymakers should establish and enforce comprehensive ethical guidelines, regulatory standards, and accountability frameworks governing the development and use of AI technologies. Such frameworks should emphasise data protection, fairness, transparency, user rights, and mechanisms for addressing AI-related errors or biases. Strong governance structures can reassure users that AI systems operate within safe and ethical boundaries, thereby fostering trust and increasing public acceptance of AI applications across various sectors.

## Data Availability

All data produced in the present study are available upon reasonable request to the authors.

